# An explorative analysis of plasma biomarkers associated with cerebral amyloid angiopathy

**DOI:** 10.1101/2025.09.03.25334994

**Authors:** Ersin Ersözlü, François Meyer, Lukas Preis, Orkan Arslan, Daria Gref, Louise von Droste, Julian Hellmann-Regen, the Alzheimer’s Disease Neuroimaging Initiative

## Abstract

**Background:** Cerebral amyloid angiopathy (CAA) remains diagnostically challenging, particularly in asymptomatic individuals. While CAA often co-exists with Alzheimer’s disease (AD), it may even have a direct impact on AD pathophysiology and the cognitive decline within the clinical course of AD. While fluid biomarkers are well-established for AD pathology, reliable markers to improve the characterization of CAA are lacking.

**Methods:** We analyzed two subsets of participants from the Alzheimer’s Disease Neuroimaging Initiative: one with available T2*-weighted gradient echo magnetic resonance imaging (MRI) (n=21) and another with postmortem neuropathological data (n=24), all with available plasma biomarkers from a 145-analyte multiplex immunoassay panel. We defined CAA as two or more lobar microbleeds in MRI or moderate to severe neocortical amyloid angiopathy in neuropathological examination. Plasma analytes were assessed twice per subject, one year apart, with the earlier sample obtained up to 6.6 years prior to either the first MRI or neuropathological examination. Non-parametric correlation and receiver operating characteristic curves were mainly reported.

**Results:** In both cohorts, various markers related to inflammation, lipid metabolism, and cell adhesion were associated with CAA proxy measures. Specifically, both increased (Osteopontin, VCAM-1) and decreased (vitronectin or endothelial growth factor) biomarker levels were associated with MBs, while increased apolipoproteins (ApoAII, ApoCI and ApoCIII, ApoE and clusterin) and decreased AXL were associated with CAA severity in neuropathology. Ratios between inversely associated markers enhanced correlation strength and discriminated CAA status.

**Conclusions:** Several candidate plasma biomarkers of CAA were identified in individuals with either MRI or neuropathological indicators of CAA.

## Background

Cerebral amyloid angiopathy (CAA) is a cerebrovascular disorder caused by the buildup of β-amyloid (Aβ) in the walls of leptomeningeal and cortical blood vessels. CAA affects 25% of the general elderly population^1^ and can clinically manifest with intracerebral hemorrhage, temporary disturbances in cortical functions (such as motor, somatosensory, visual, or language functions), and progressive cognitive decline^2^. CAA shares key pathological characteristics with Alzheimer’s disease (AD), including the accumulation of Aβ-peptides. Notably, in AD, Aβ plaques primarily accumulate in the brain parenchyma adjacent to neurons, whereas CAA involves Aβ buildup outside the brain in cerebral blood vessels^3,4^. In around 50% of cases, CAA co-occurs with AD^1^.

Previously, diagnosing CAA relied on postmortem brain tissue examination. The introduction of the Boston Criteria enabled the diagnosis of in vivo CAA, defining “possible” or “probable” CAA by using mainly clinical and magnetic resonance imaging (MRI) findings such as lobar microbleeds (MBs)^5^. The Boston Criteria v2.0 has extended the criteria notably with clinical features and non-hemorrhagic findings^6^. However, when applied to asymptomatic patients or patients with cognitive impairment, the v.2.0 criteria revealed a 28.6% sensitivity and 65.3% specificity for “probable CAA” only^7^. This can be considered rather low, compared to diagnostic performances in the field of AD, where fluid biomarkers in cerebrospinal fluid (CSF) and plasma have evolved into reliable diagnostic tests^8^. A limited number of studies have addressed the unmet need for accurate diagnostic approaches to CAA and found that CAA may be differentiated from AD by lower CSF Aβ40 and Aβ42 levels, while CAA-related inflammation has been linked to activation of microglia and T cells as well as increased CSF tau concentrations^9,10^.

Given the heterogeneous clinical manifestation and the limited diagnostic accuracy of present diagnostic strategies, we aimed to conduct an exploratory analysis of candidate plasma markers of CAA by using MRI and neuropathological indicators of CAA.

## Methods

### Participants

Data used in the preparation of this article were obtained from the Alzheimer’s Disease Neuroimaging Initiative (ADNI) database (adni.loni.usc.edu). Participants included in our analyses were recruited for the prospective the Alzheimer’s Disease Neuroimaging Initiative (ADNI)-1 convenience cohort (National Clinical Trial (NCT): 00106899), while MRI data were acquired during follow-up visits within ADNI-go, ADNI-2 and ADNI-3 convenience cohorts (NCT00106899, NCT01231971 and NCT02854033).

A summary of inclusion and exclusion criteria and detailed characteristics of the included study sample are provided in the supplementary material. In short, we included participants with available plasma biomarkers and either with available visual assessments of MBs in T2*-GRE MRI (cohort-MRI, n=21) or postmortem neuropathological assessment (cohort-NP, n=24) **(Supplementary Figure 1)**.

### Study design

For the primary analysis, we selected a time point (T0) when all multiplex panel measurements (MPM) were accessible. Using a longitudinal approach, we also investigated the associations of a second MPM after one year (T1) with radiologic and neuropathological assessments for lobar MBs (**Figure 1A**) and neuropathological findings indicative of CAA (**Figure 1B**), respectively.

**Figure 1.**
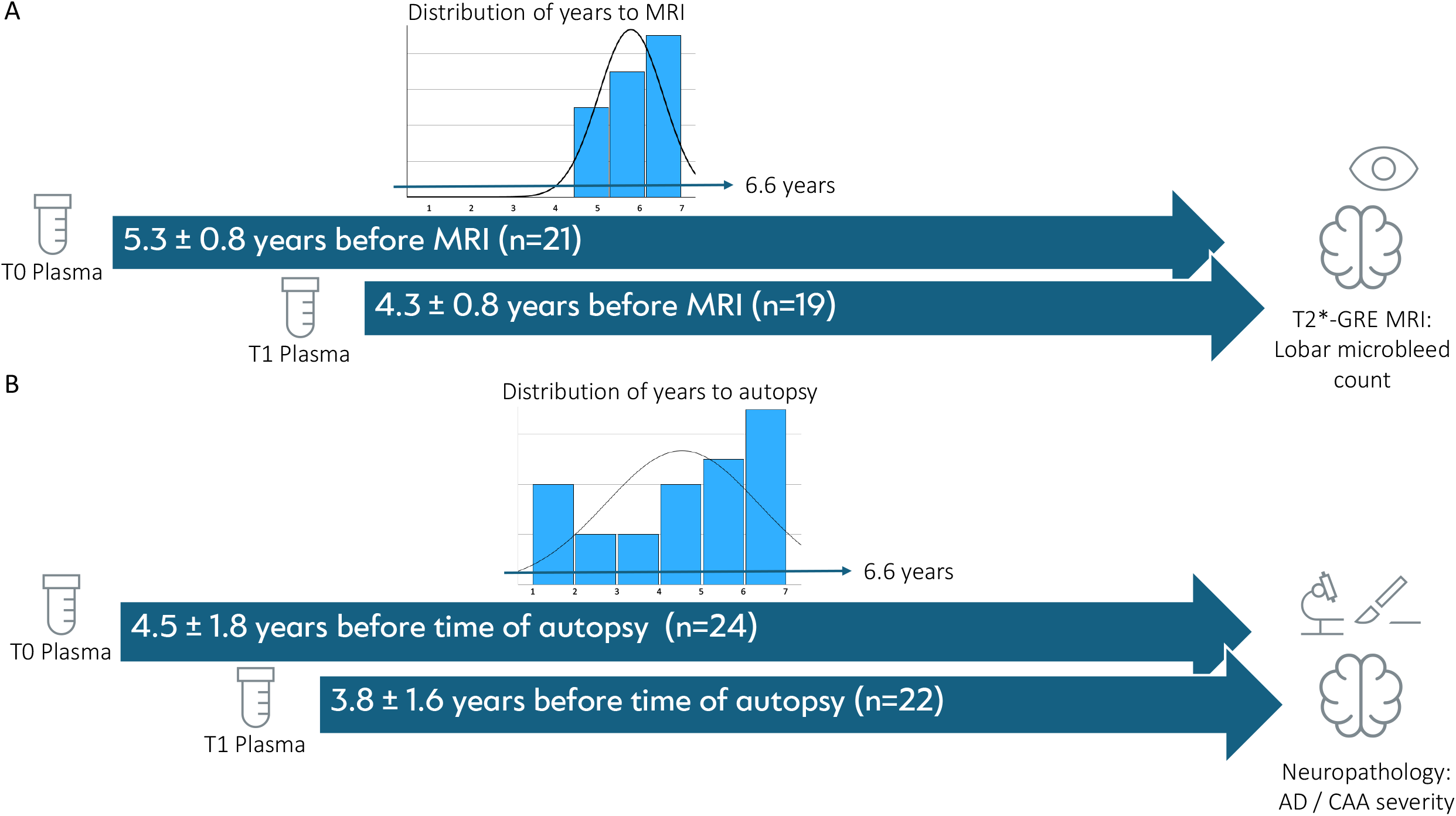
Graphical summary of the study design. Abbreviations: AD, Alzheimer’s disease; CAA, cerebral amyloid angiopathy.

### Fluid biomarkers

For the plasma measurements, we included all available analytes (n=145) in the multiplex immunoassay panel, consisting of proteins related to cancer, cardiovascular disease, metabolic disorders, inflammation, and AD— see **Supplementary Table 1**. Notably, plasma analytes were measured at time points T0 and T1, with a one-year interval. Summarized descriptions of measurements of plasma and CSF markers are presented in the supplementary material. The plasma samples were analyzed by using the Luminex xMAP (Multi-Analyte Profiling) technology, while the CSF samples were analyzed using the electrochemiluminescence immunoassay (ECLIA) Elecsys on a fully automated Elecsys cobas e instrument (Roche Diagnostics GmbH, Penzberg, Germany). The cut-off value of 0.1 was defined for p-tau181p/Aβ1–42 as reported elsewhere^11^.

### Cranial MRI Acquisition and Analysis

The imaging protocol includes multicentric 3T MRI examinations including GE, Siemens, and Philips MRI Systems, among others including T2*-GRE sequences (repetition time (TR) 650 ms, echo time (TE) 20 ms, flip angle (FA) 20 degrees) and T1-MPRAGE (TR 2300 ms, TE 2.95 ms, FA 9 degrees) sequences. T2* imaging enables in vivo detection of microhemorrhage-derived hemosiderin deposits as a hypointense signal.

Detailed descriptions of MB detection and atlas assignment methods have been reported in the ADNI Methods document, “Mayo (Jack Lab) – ADNI MRI MCH [ADNIGO,2,3], version 2021-12-13” by Clifford R. Jack, Jr., M.D., from the Mayo Clinic in Rochester, MN, available at http://adni.loni.usc.edu/. A summary of image acquisition parameters, as well as quality and radiological visual assessments, is also provided in the supplementary material. Notably, the participants with two or more lobar microbleeds have been classified as CAA^6^.

### Neuropathological Assessment

Pathological lesions within the brain were assessed during autopsy using established neuropathologic diagnostic criteria. Detailed descriptions of neuropathological assessments have been reported in the ADNI Methods document, “ADNI_Neuropathology_Core_Methods_FINAL_20221114”, available at http://adni.loni.usc.edu/, and are presented in summary in the supplementary material.

The central neuropathological severity scales for AD (Variable NPADNC) and CAA (Variable NPAMY) have been included. We included the semi-quantitative (None, low, intermediate, high) assessment for AD using the Alzheimer’s Disease – NIA-AA Neuropathological Change level^12^. Likewise, the semi-quantitative assessment of overall neocortical amyloid angiopathy refers to the global CAA according to the following scale that refers to the global CAA burden: 0 — None: Absent 1 — Mild: Scattered positivity in parenchymal and/or leptomeningeal vessels, possibly in only one brain area 2 — Moderate: Intense positivity in many parenchymal and/or leptomeningeal vessels 3 — Severe: Widespread (more than one brain area) intensive positivity in parenchymal and leptomeningeal vessels.

Importantly, we classified subjects with ‘Moderate’ or ‘Severe’ CAA burden as CAA positive, based on the previous literature suggesting its clinical importance^13^.

### Statistics

All statistical analyses and graphical representations were carried out using SPSS® Statistics (IBM®, v29). Likewise, we generated the scatter plots and receiver operating characteristic (ROC) curves in SPSS®. The sample size was not predetermined by statistical methods; instead, all available data were included in the analysis.

In summary, group differences at baseline were assessed using Mann-Whitney U and chi-square tests. Nonparametric correlations were assessed with Spearman’s rank correlation. The results of the subsequent analyses were then corrected for multiple testing by using false discovery rate (FDR) with α<0.05. ROC analyses were conducted to evaluate the discriminatory performance of biomarker ratios for clinical outcome groups, reporting mainly area under the curves (AUCs). The detailed descriptions of the statistical methods are presented in the supplementary material.

## Results

The CAA groups in cohort-MRI and cohort-NP did not differ in demographic, genetic, and clinical features, except clinical diagnosis (**Supplementary Table 2**). Additionally, we presented the group characteristics of subgroups divided by CAA status within both cohorts (**Supplementary Table 3**), revealing no differences except that Aβ42 was lowered in the CAA+ group in cohort-NP.

### Linking Plasma Biomarkers to CAA Proxy Measures

The correlation analyses revealed several positive and negative associations. Positive associations included adhesion (VCAM-1), vascular (Angiopoietin-2, RAGE), and immune response/inflammation (Osteopontin, FASLG, NGAL) markers correlating with MB count and several apolipoproteins (ApoAII, ApoCI, ApoCIII, ApoE and CLU), ComFH, TTR and Leptin correlating with neuropathological CAA severity (**Supplementary Table 4**). Due to inverse relationships among analytes and to address physiological limitations in measuring plasma analytes we propose several ratios that partly strengthened the correlations (VCAM1/vitronectin, FASLG/EGF Ratio, NGAL/EGF Ratio, and RAGE/EGF in cohort-MRI; ApoAII/AXL, ApoCI/AXL, ApoCIII/AXL, ApoE/AXL, CLU/AXL, ComFH/AXL, TTR/AXL, and Leptin/AXL in cohort-NP) (**Table 1, Supplementary Figure 2)**.

**Figure 2.**
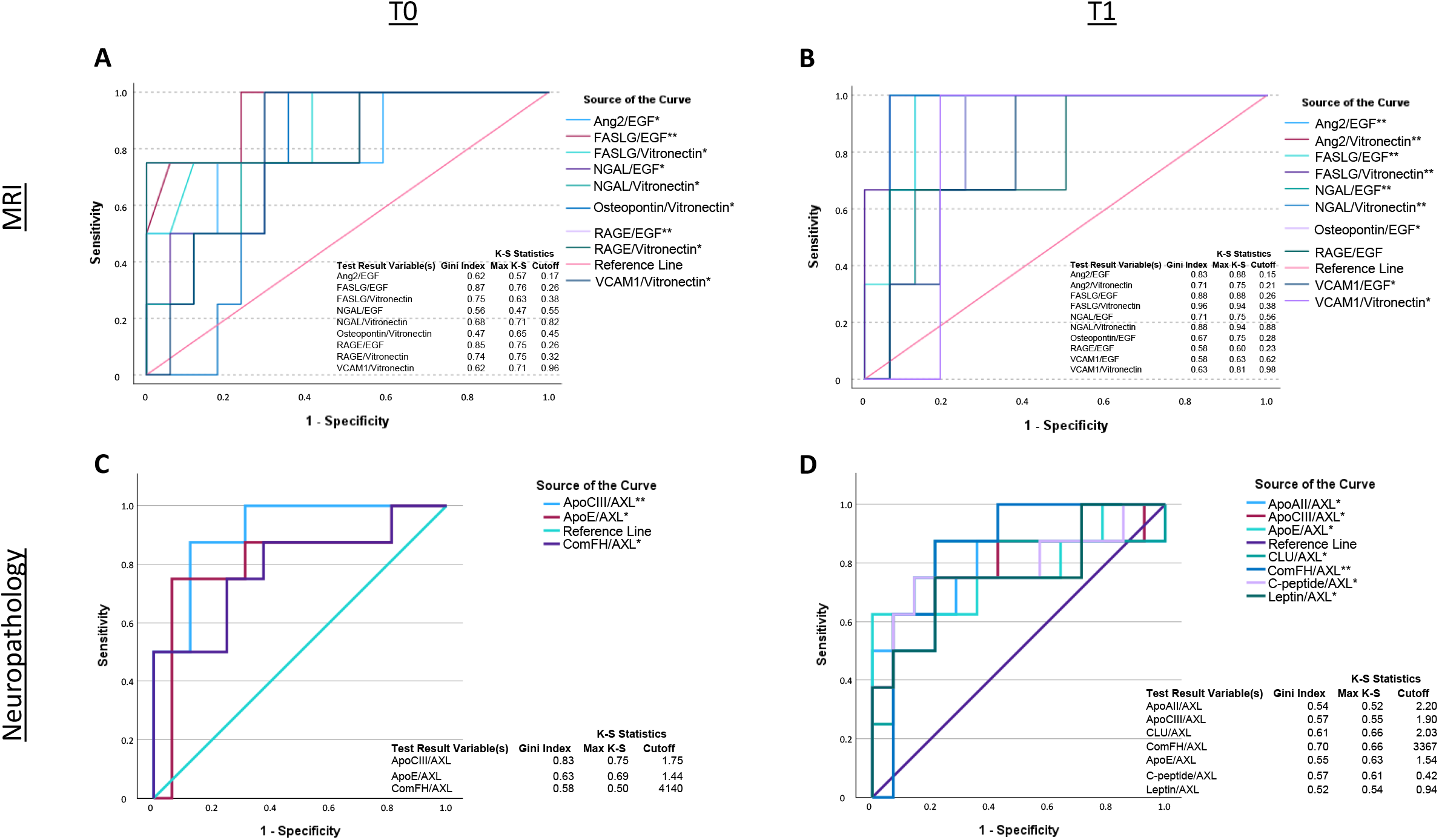
ROC curves of ratios of analytes in the differentiation of CAA among the study groups and time points. *False discovery rate (FDR) corrected-p<0.05; **FDR-p<0.001. Abbreviations: CAA, cerebral amyloid angiopathy; Ang2, Angiopoietin-2; Epidermal Growth Factor Receptor; NGAL, Neutrophil Gelatinase-Associated Lipocalin; RAGE, Receptor for Advanced Glycation End; VCAM1, Vascular Cell Adhesion Molecule-1; Apo, Apolipoprotein; CLU, Clusterin; ComFH, Complement Factor H.

**Table 1.**
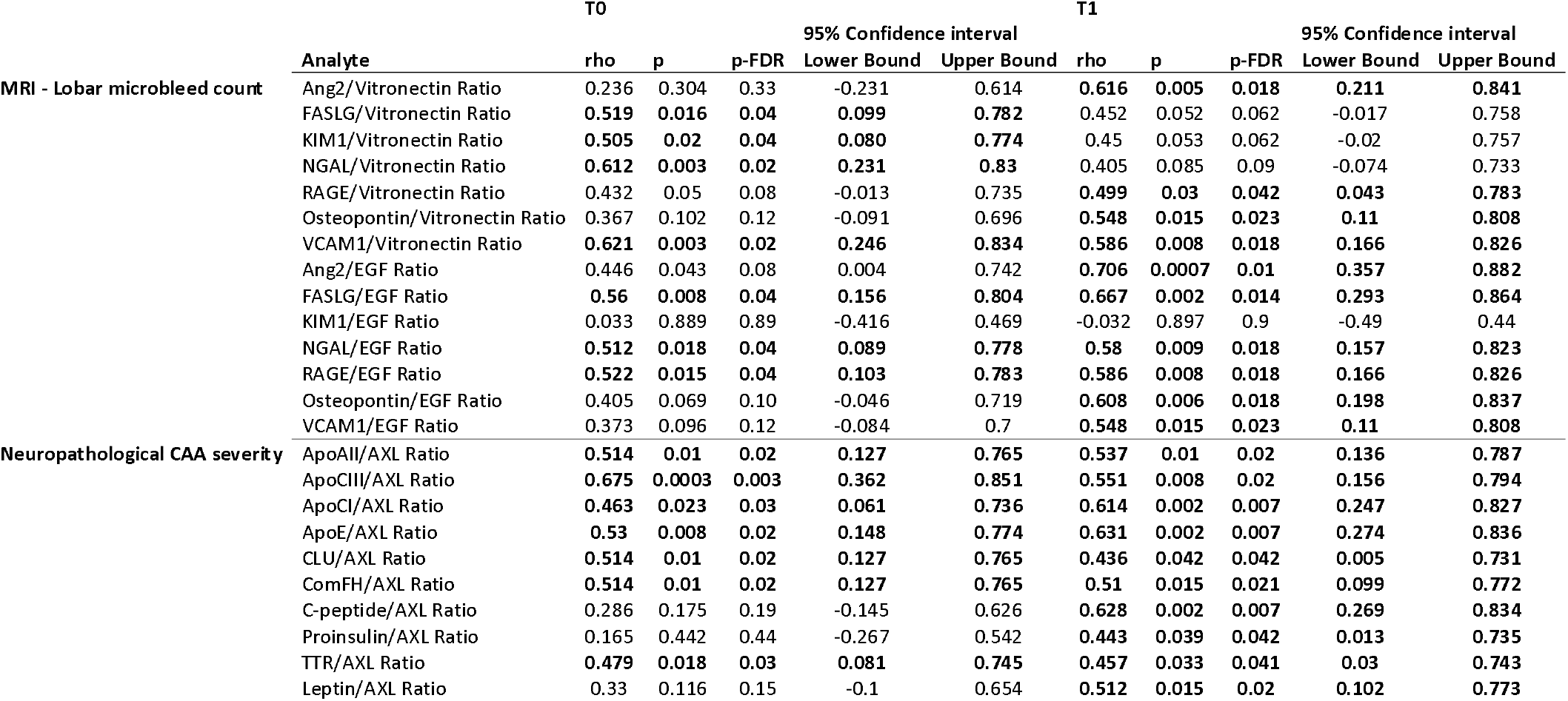
Correlations of lobar microbleed count and neuropathological cerebral amyloid angiopathy severity score with plasma analytes. Only results with p<0.05 in both time points are presented. Results with false discovery rate (FDR) corrected p <0.05 are marked in bold. Abbreviations: Ang2, Angiopoietin-2; FASLG, FAS Ligand Receptor; KIM1, Kidney Injury Molecule-1; NGAL, Neutrophil Gelatinase-Associated Lipocalin; RAGE, Receptor for advanced glycosylation end; VCAM1, Vascular Cell Adhesion Molecule-1; EGF, Epidermal Growth Factor Receptor; Apo, Apolipoprotein; CLU, Clusterin; ComFH, Complement Factor H; TTR, Transthyretin.

### Discrimination of CAA Status Based on Plasma Biomarker Levels

To further test whether the ratios can discriminate CAA status separately in both cohorts, binarized CAA status was discriminated by ratios of data-driven selected plasma markers with moderate to high discrimination power AUCs 0.74-0.98 for cohort-MRI and AUCs 0.76-0.91 for cohort-NP) (**Figure 2, Supplementary Table 5**). The highest three AUC values have been found for FASLG/EGF (0.93, 95% CI 0.81-1.06), RAGE/EGF (0.93, 95% CI 0.78-1.07) and FASLG/Vitronectin (0.88, 95% CI 0.68-1.07) at T0 (**Figure 2A**) and for FASLG/Vitronectin (0.98, 95% CI 0.92-1.04), NGAL/Vitronectin (0.94, 95% CI 0.82-1.06) and FASLG/EGF (0.94, 95% CI 0.83-1.05) at T1 (**Figure 2B**) in cohort-MRI. The highest three AUC values were revealed for ApoCIII/AXL (0.91, 95% CI, 0.8-1.03), ApoE/AXL (0.81, 95% CI, 0.6-1.032) and ComFH/AXL (0.79, 95% CI, 0.58-1) at T0 (**Figure 2C**) and for ComFH/AXL (0.85, 95% CI 0.68-1.02), CLU/AXL (0.8, 95% CI 0.57-1.04) and ApoCIII (0.79, 95% CI 0.56-1.01) at T1 (**Figure 2D**) in cohort-NP.

### Trajectories of lobar microbleeds and their correlation with plasma analyte ratios

To evaluate the correlations between plasma analyte ratios and longitudinal trajectories of MBs, we calculated annual change rate of MBs. Here, we observed that Ang2/EGF positively correlated with change in MBs at both time points, while not remaining significant following the FDR correction. Considering further uncorrected results, FASLG/vitronectin and Osteopontin/vitronectin revealed positive correlations at T1 and ApoE/AXL a negative correlation at T2 (**Supplementary Table 6, Supplementary Figure 3**).

## Discussion

Examining the associations of ante-mortem and post-mortem indicators of CAA with plasma marker levels separately, we identified several plasma biomarkers related to immune activation, inflammation, cell adhesion, apoptosis, and glycosylation as candidate biomarkers of CAA. However, the identified candidate markers differed depending on the modality used to define CAA (MRI or neuropathology) and are also likely to reflect processes such as aging or non-CAA pathologies.

Among other markers, higher levels of FASLG/EGF and FASLG/vitronectin ratios have been observed in participants with more than two lobar microbleeds at both time points, as well as longitudinally with changes in lobar microbleed count in a U-shaped manner (uncorrected). FASLG, also known as CD95L, is associated with astrocyte reactivation and cell apoptosis, while being reported in neuroinflammatory and neurodegenerative diseases as well as ischemic stroke^14,15^. Moreover, FASLG upregulation has been observed in in vitro AD models and contributed to Aβ-mediated neuronal cell death^16^.

Further elevation was found for osteopontin/EGF and osteopontin/vitronectin in relation to higher microbleed counts. Higher osteopontin/vitronectin levels also predicted a longitudinal increase in lobar microbleed count, although this did not survive correction for multiple testing. Osteopontin is an extracellular phosphoprotein and potentially related to pathological processes in AD and CAA, as well as vascular dementia^17,18^. The mechanisms of osteopontin upregulation in neurodegenerative diseases might include neuroinflammatory or compensatory response^17^.

We further identified several glycoproteins, such as clusterin, which is multifunctional and has been linked to the pathophysiology of cerebral amyloidosis in both AD^19^ and CAA^20^, where the authors also identified upregulation of further proteins, including ApoE and vitronectin, by examining biopsied leptomeningeal and cortical vessels of patients with CAA^20^. More interestingly, the authors also found an in vitro model of CAA in which physiological concentrations of ApoE and clusterin might inhibit Aβ amyloid deposition in CAA^20^. Considering the mixed results, one possible interpretation of our results is that ApoE and clusterin might be elevated in response to emerging CAA pathology, as plasma levels have been measured many years before the neuropathological assessment. In contrast, our results in the MRI cohort revealed a negative association of vitronection with MBs. We argue that the difference might have occurred due to the difference in modality - MRI versus pathological assessment.

Microglial activity is involved in the pathophysiological pathways leading to CAA pathology, and TAM receptors mediate the glial response to cerebral and vascular Aβ plaques^10,21^. With this in mind, regarding the stronger associations of proportions built with AXL compared to single analytes such as proinflammatory ApoCIII^22^, a reduced capacity of microglial response can be argued alongside the upregulated markers of immunity and inflammation.

Confirmatory studies are crucial to address possible confounding factors and temporal relationships. Importantly, the study group characteristics revealed unremarkable differences between CAA and non-CAA groups. Further limitations include the partly applied Boston criteria v2.0, which allows for defining probable CAA with only one MB when typical non-hemorrhagic findings are present. This decision was made to define probable CAA cases more homogeneously, considering the limited sample size. With this in mind, the markers predicting MBs might be nonspecific to CAA and could offer screening of MBs resulting from various etiologies.

While these exploratory results certainly need replication in another CAA cohort, our findings might contribute to a more precise diagnosis of CAA and, regarding the participants with concomitant AD pathology, also to estimate probabilities of response and risk related to future combination strategies of new pharmacological approaches^23^, especially anti-amyloid antibodies and amyloid-related imaging abnormalities^10^.

## Data Availability

The data supporting this study's findings are openly available in the ADNI database at www.loni.ucla.edu/ADNI.

## Acknowledgement

Data collection and sharing for the Alzheimer’s Disease Neuroimaging Initiative (ADNI) is funded by the National Institute on Aging (National Institutes of Health Grant U19AG024904). The grantee organization is the Northern California Institute for Research and Education. In the past, ADNI has also received funding from the National Institute of Biomedical Imaging and Bioengineering, the Canadian Institutes of Health Research, and private sector contributions through the Foundation for the National Institutes of Health (FNIH) including generous contributions from the following: AbbVie, Alzheimer’s Association; Alzheimer’s Drug Discovery Foundation; Araclon Biotech; BioClinica, Inc.; Biogen; BristolMyers Squibb Company; CereSpir, Inc.; Cogstate; Eisai Inc.; Elan Pharmaceuticals, Inc.; Eli Lilly and Company; EuroImmun; F. Hoffmann-La Roche Ltd and its affiliated company Genentech, Inc.; Fujirebio; GE Healthcare; IXICO Ltd.; Janssen Alzheimer Immunotherapy Research & Development, LLC.; Johnson & Johnson Pharmaceutical Research & Development LLC.; Lumosity; Lundbeck; Merck & Co., Inc.; Meso Scale Diagnostics, LLC.; NeuroRx Research; Neurotrack Technologies; Novartis Pharmaceuticals Corporation; Pfizer Inc.; Piramal Imaging; Servier; Takeda Pharmaceutical Company; and Transition Therapeutics.

## Disclosures

The authors report no disclosures related to the study.

## Notes

### Competing Interest Statement

The authors have declared no competing interest.

### Author Declarations

Data used in the preparation of this article were obtained from the Alzheimer's Disease Neuroimaging Initiative (ADNI) database (adni.loni.usc.edu).

