## Supplementary material for "An explorative analysis of plasma biomarkers associated with cerebral amyloid angiopathy"

*Data used in preparation of this article were obtained from the Alzheimer’s Disease Neuroimaging Initiative (ADNI) database (adni.loni.usc.edu). As such, the investigators within the ADNI contributed to the design and implementation of ADNI and/or provided data but did not participate in analysis or writing of this report. A complete listing of ADNI investigators can be found at: <http://adni.loni.usc.edu/wp-content/uploads/how_to_apply/ADNI_Acknowledgement_List.pdf>

Methods

Participants

Data used in the preparation of this article were obtained from the Alzheimer's Disease Neuroimaging Initiative (ADNI) database (adni.loni.usc.edu). The ADNI was launched in 2003 as a public-private partnership, led by Principal Investigator Michael W. Weiner, MD. The original goal of ADNI was to test whether serial magnetic resonance imaging (MRI), positron emission tomography (PET), other biological markers, and clinical and neuropsychological assessment can be combined to measure the progression of mild cognitive impairment (MCI) and early Alzheimer's disease (AD). The current goals include validating biomarkers for clinical trials, improving the generalizability of ADNI data by increasing diversity in the participant cohort, and providing data concerning the diagnosis and progression of Alzheimer’s disease to the scientific community. For up-to-date information, see adni.loni.usc.edu.

In ADNI study, participants who were between 55 and 90 (inclusive) years of age, had a study partner able to provide an independent evaluation of functioning, and spoke either English or Spanish were enrolled. The Mini-Mental State Examination, Clinical Dementia Rating, and adjusted Wechsler Memory Scale - Logical Memory II have been considered for the clinical diagnoses, together with NINCDS/ADRDA criteria for probable AD.

In the current study, we analyzed the data of a total of 45 participants with available plasma biomarkers from a multiplex immunoassay panel and available visual assessments of microbleeds (MBs) in T2*-GRE MRI (Cohort-MRI, n=21) and had postmortem neuropathological assessment (cohort-NP, n=24). Of note, to match the time intervals between GRE and NP groups, we limited the highest time difference to equal or less than 6.6 years, based on the highest time difference in the Cohort-MRI. Furthermore, as two participants had both available data for MRI and NP, they were included only in the MRI cohort due to the smaller size of the MRI cohort.

The definitions of clinical variables regarding the cardiovascular risk factors have been described in in ADNI documentation “ADSP Phenotype Harmonization Consortium – Derivation of Harmonized Cardiovascular Risk Factors” that is available at <https://adni.loni.usc.edu/data-samples/access-data/>. Notably, the binary variables have been recorded from as either “Ever Had” or “Never Had” for the heart disease, stroke, hypertension and diabetes. The smoking and hypertensive medication statuses were recorded regarding the current state.

Fluid biomarkers

The detailed methods of plasma measurements, including quality assessment, are reported in described in ADNI documentation “Biomarkers Consortium Plasma Proteomics Data Primer 02Aug2013 FINAL” that is available at <https://adni.loni.usc.edu/data-samples/access-data/>. Notably, the documentation reported that plasma samples were obtained in the morning following an overnight fast, and the time from collection to freezing was within 120 minutes for the majority of samples. The quality-controlled and cleaned dataset titled “adni_plasma_qc_multiplex_11Nov2010” has been used.

CSF biomarker concentrations for Aβ1–42, phosphorylated-tau181 (p-tau181), and total-tau (t-tau) were available at timepoint T0 for n = 17 (81%) and 18 (75%) for MRI and neuropathology cohorts, respectively. Aliquoted samples were analyzed using the electrochemiluminescence immunoassay (ECLIA) Elecsys on a fully automated Elecsys cobas e instrument (Roche Diagnostics GmbH, Penzberg, Germany) as fully described in ADNI documentation “ADNI3: Batch analysis of Aβ1-42, t-tau and p-tau181 in ADNI1 and ADNIGO/2 CSF using the fully automated Roche Elecsys and cobas e immunoassay analyzer system. 2017.” which is available at <https://adni.loni.usc.edu/data-samples/access-data/>.

Cranial MRI Acquisition and Analysis

Experienced raters manually counted MB in T2* native space images. These assessments were conducted within a longitudinal framework at the Aging and Dementia Imaging Research Laboratory at the Mayo Clinic. Each MB was spatially assigned using a T1-MPRAGE image, mapped onto a 35-region atlas from the Mayo Clinic Adult Lifespan Template, which is accessible via NITRC (<https://www.nitrc.org/projects/mcalt>). To facilitate automatic atlas region assignment, the T1-MPRAGE image was affine-registered to the T2*-GRE image. The localization and classification of each MB finding—including its assigned atlas region and tissue probability (gray matter, white matter, or CSF)—were documented. The central visual ratings were performed by five Medical Imaging Analysts with specialized training from the Mayo Clinic’s Department of Radiology, subsequently reviewed by a board-certified neuroradiologist for confirmation. All MH investigators received the images anonymized and were blinded to the clinical diagnosis. Finally, we considered only the possible and definite lobar MB counts from the central visual readings by the Mayo Clinic for our analyses due to their known associations with CAA rather than non-lobar MB.

Annual change rate of lobar MBs was obtained by dividing the difference between follow-up and baseline MRI examinations by the difference in years between both examinations.

Statistics

Group differences at T0 were compared with the Mann-Whitney U test for continuous variables and the chi-square test for binary variables, as appropriate.

Spearman’s rank-order correlation coefficients of MB count and NPAMY were computed to assess their nonparametric correlations with the above-mentioned panel of plasma biomarkers. Correlation analyses were conducted using 95% confidence intervals, estimating the variance suggested by Fieller, Hartley and Pearson. Correlation results were interpreted without significance thresholding to allow for exploratory analysis, while results are considered significant when two-tailed p<0.05.

Then we derived several biomarker ratios to account for relative expression levels, regarding their inverse correlations. Specifically, ratios of selected biomarkers with positive associations with CAA proxy measures (e.g., Angiopoietin-2, FASLG receptor, KIM-1, NGAL, RAGE, Osteopontin, and VCAM-1) to Vitronectin or EGFR were calculated. Furthermore, ratios of apolipoproteins and adhesion and metabolic markers (e.g., ApoC-III, ApoE, Clusterin, C-peptide, Leptin, Proinsulin, and Transthyretin) to soluble AXL were computed. These ratios were included in subsequent nonparametric correlation analysis with MB count and NPAMY, separately. Here, we applied False Discovery Rate (FDR) correction for multiple testing with α<0.05.

We tested the nonparametric correlations between the plasma analyte ratios and the annual change rate of lobar MBs. Again, we applied FDR correction for multiple testing with α<0.05.

Receiver Operating Characteristic (ROC) analyses were performed to evaluate the diagnostic performance of the various above-mentioned biomarker ratios in distinguishing between clinical outcome groups. Ratios of biomarkers were analyzed for their ability to differentiate participants with either (1) two or more MB or (2) moderate or severe NPAMY. Moreover, ROC analyses were carried out with the assumption of a nonparametric distribution of test variables. Test positivity was defined in the direction of larger biomarker values. For each variable, the area under the ROC curve (AUC) was calculated along with standard errors and 95% confidence intervals. ROC curves and classification performance metrics were generated to assess model quality. Again, we applied FDR correction for multiple testing with α<0.05.

Supplementary Table 1. List of the included quality-assessment passed plasma analytes.

**Analytes**

Adiponectin

Agouti-Related Protein (AGRP)

Alpha-1-Antichymotrypsin (AACT)

Alpha-1-Antitrypsin (AAT)

Alpha-1-Microglobulin (A1Micro)

Alpha-2-Macroglobulin (A2Macro)

Alpha-Fetoprotein (AFP)

Angiopoietin-2 (ANG-2)

Angiotensin-Converting Enzyme (ACE)

Angiotensinogen

Apolipoprotein A-I (ApoAI)

Apolipoprotein A-II (ApoAII)

Apolipoprotein A-IV (ApoAIV)

Apolipoprotein B (ApoB)

Apolipoprotein C-I (ApoCI)

Apolipoprotein C-III (ApoCIII)

Apolipoprotein D (ApoD)

Apolipoprotein E (ApoE)

Apolipoprotein H (ApoH)

Apolipoprotein(a) (Lp(a))

AXL Receptor Tyrosine Kinase (AXL)

B Lymphocyte Chemoattractant (BLC)

Beta-2-Microglobulin (B2M)

Betacellulin (BTC)

Bone Morphogenetic Protein 6 (BMP-6)

Brain Natriuretic Peptide (BNP)

Brain-Derived Neurotrophic Factor (BDNF)

Calcitonin

Cancer Antigen 19-9 (CA-19-9)

Carcinoembryonic Antigen (CEA)

CD 40 antigen (CD40)

CD40 Ligand (CD40-L)

CD5 (CD5L)

Chemokine CC-4 (HCC-4)

Chromogranin-A (CgA)

Ciliary Neurotrophic Factor (CNTF)

Clusterin (CLU)

Complement C3 (C3)

Complement Factor H (ComFH)

Cortisol (Cortisol)

C-peptide

C-Reactive Protein (CRP)

Creatine Kinase-MB (CK-MB)

Cystatin-C

Eotaxin-1

Eotaxin-3

Epidermal Growth Factor (EGF)

Epithelial-Derived Neutrophil-Activating Peptide-78

E-Selectin

Factor VII

Fas Ligand (FasL)

FASLG Receptor (FASLG)

Fatty Acid-Binding Protein- heart (FABP-h)

Ferritin (FRTN)

Fetuin-A

Fibrinogen

Fibroblast Growth Factor 4 (FGF-4)

Follicle-Stimulating Hormone (FSH)

Glutathione S-Transferase alpha (GST-alpha)

Growth Hormone (GH)

Growth-Regulated alpha protein (GRO-alpha)

Haptoglobin

Heparin-Binding EGF-Like Growth Factor

Hepatocyte Growth Factor (HGF)

Immunoglobulin A (IgA)

Immunoglobulin E (IgE)

Immunoglobulin M (IGM)

Insulin

Insulin-like Growth Factor-Binding Protein

Intercellular Adhesion Molecule 1 (ICAM-1)

Interferon gamma Induced Protein 10 (IP-10)

Interleukin-13 (IL-13)

Interleukin-16 (IL-16)

Interleukin-18 (IL-18)

Interleukin-3 (IL-3)

Interleukin-6 receptor (IL-6r)

Interleukin-8 (IL-8)

Kidney Injury Molecule-1 (KIM-1)

Leptin

Luteinizing Hormone (LH)

Macrophage Colony-Stimulating Factor 1 (MCP-1)

Macrophage Inflammatory Protein-1 alpha (MIP-1-alpha)

Macrophage Inflammatory Protein-1 beta (MIP-1-beta)

Macrophage Inflammatory Protein-3 alpha (MIP-3-alpha)

Macrophage Migration Inhibitory Factor (MMIF)

Macrophage-Derived Chemokine (MDC)

Matrix Metalloproteinase-1 (MMP-1)

Matrix Metalloproteinase-10 (MMP-10)

Matrix Metalloproteinase-2 (MMP-2)

Matrix Metalloproteinase-7 (MMP-7)

Matrix Metalloproteinase-9 (MMP-9)

Matrix Metalloproteinase-9- total

Monocyte Chemotactic Protein 1 (MCP-1)

Monocyte Chemotactic Protein 2 (MCP-2)

Monocyte Chemotactic Protein 3 (MCP-3)

Monocyte Chemotactic Protein 4 (MCP-4)

Monokine Induced by Gamma Interferon

Myeloid Progenitor Inhibitory Factor 1

Myeloperoxidase (MPO)

Myoglobin

Neuronal Cell Adhesion Molecule (Nr-CAM)

Neutrophil Gelatinase-Associated Lipocalin (NGAL)

Osteopontin

Pancreatic Polypeptide (PPP)

Peptide YY (PYY)

Placenta Growth Factor (PLGF)

Plasminogen Activator Inhibitor 1 (PAI-1)

Platelet-Derived Growth Factor BB (PDGF-BB)

Pregnancy-Associated Plasma Protein A (PAPPA)

Proinsulin-Intact (pM)

Proinsulin-Total (pM)

Prolactin (PRL)

Prostatic Acid Phosphatase (PAP)

Pulmonary and Activation-Regulated Chemokine (PARC)

Receptor for advanced glycosylation end (RAGE)

Resistin

Serotransferrin (Transferrin)

Serum Amyloid P-Component (SAP)

Serum Glutamic Oxaloacetic Transaminase

Sex Hormone-Binding Globulin (SHBG)

Sortilin

Stem Cell Factor (SCF)

Superoxide Dismutase 1- Soluble (SOD-1)

T Lymphocyte-Secreted Protein I-309 (I-309)

Tamm-Horsfall Urinary Glycoprotein (THP)

T-Cell-Specific Protein RANTES (RANTES)

Tenascin-C (TN-C)

Testosterone-Total

Thrombomodulin (TM)

Thrombopoietin

Thrombospondin-1

Thymus-Expressed Chemokine (TECK)

Thyroid-Stimulating Hormone (TSH)

Thyroxine-Binding Globulin (TBG)

Tissue Inhibitor of Metalloproteinases 1

TNF-Related Apoptosis-Inducing Ligand Re

Transthyretin (TTR)

Trefoil Factor 3 (TFF3)

Tumor Necrosis Factor alpha (TNF-alpha)

Tumor Necrosis Factor Receptor-Like 2 (TNF Receptor-Like 2)

Vascular Cell Adhesion Molecule-1 (VCAM-1)

Vascular Endothelial Growth Factor (VEGF)

Vitamin K-Dependent Protein S (VKDPS)

Vitronectin

von Willebrand Factor (vWF)

Supplementary Table 2. Group characteristics among cohorts. Mean (± standard deviations) are shown for continuous variables and number (percent) are presented for categorical or ordinal variables if not mentioned otherwise.

*Abbreviations: MRI, magnetic resonance imaging; NP, neuropathology; ApoE, ApolipoproteinE; NA, not applicable.*

|  | Cohort-MRI (N=21) | Cohort-NP (N=24) | p |
| --- | --- | --- | --- |
| Age | 72 ± 9 | 77 ± 7 | 0.08* |
| Female | 6 (27%) | 4 (16.7%) | 0.39♰ |
| Educational years | 16 ± 3 | 16 ± 2 | 0.65* |
| Clinical Diagnosis |  |  | 0.004♰ |
| Cognitively normal | 4 (18%) | 0 (0%) |  |
| Mild cognitive impairment | 17 (82%) | 18 (75%) |  |
| Dementia | 0 (0%) | 6 (25%) |  |
| *ApoE ε4* count |  |  | 0.37♰ |
| 0 | 13 (59%) | 10 (41.7%) |  |
| 1 | 5 (23%) | 10 (41.7%) |  |
| 2 | 3 (14%) | 4 (16.7%) |  |
| CSF Phospho-tau/Beta-Amyloid 1-42 Ratio positive status | 7 (41.2%‡) | 13 (72%‡) | 0.06♰ |
| Time: Plasma to first MRI | 5.3 ± 0.8 | NA | - |
| Time: Plasma to time of autopsy | NA | 4.5 ± 1.8 | - |
| Lobar cerebral microbleed count, median (Range) | 0 (0-14) | NA | - |
| ≥2 lobar cerebral microbleeds | 4 (19%) | NA | - |
| Plasma panel data available at T1 | 19 (90%) | 22 (92%) | - |
| Change in lobar microbleeds, median (Range) | 0 (0-3)‡‡ | NA | - |
| Time: Baseline MRI to follow-up MRI, months | 45 ± 32‡‡ | NA | - |
| Alzheimer’s Disease – NIA-AA Neuropathological Change level |  |  | - |
| 0 (None) | NA | 1 (4.2%) |  |
| 1 (Low) | NA | 6 (25%) |  |
| 2 (Intermediate) | NA | 0 (0%) |  |
| 3 (High) | NA | 17 (70.8%) |  |
| Cerebral Amyloid Angiopathy – Density |  |  | - |
| 0 (None) | NA | 2 (8.3%) |  |
| 1 (Mild) | NA | 14 (58.3%) |  |
| 2 (Moderate) | NA | 2 (8.3%) |  |
| 3 (Severe) | NA | 6 (25%) |  |
| * Mann-Whitney U test.  ♰ Chi-square Test.  ‡ Percentage among the available cases (available in n=17 and n=18 for MRI and NP cohorts, respectively)  ‡‡ Percentage among the available cases (available in n=17) | | | |

Supplementary Table 3. Group characteristics among CAA groups in both cohorts. Mean (± standard deviations) are shown for continuous variables and number (percent) are presented for categorical or ordinal variables if not mentioned otherwise.

*Abbreviations: MRI, magnetic resonance imaging; NP, neuropathology; ApoE, ApolipoproteinE; NA, not applicable.*

|  | **Cohort-MRI (N=21)** | | | **Cohort-NP (N=24)** | | |
| --- | --- | --- | --- | --- | --- | --- |
|  | **CAA- (n=17)** | **CAA+ (n=4)** | **p** | **CAA- (n=16)** | **CAA+ (n=8)** | **p** |
| **Age** | 71 ± 10 | 75 ± 3 | 0.24* | 76.8 ± 7 | 75.9 ± 6 | 0.53* |
| **Female** | 5 (29.4%) | 1 (25%) | 0.86♰ | 1 (6.25%) | 3 (37.5%) | 0.053♰ |
| **Educational years** | 17 ± 3 | 17 ± 2 | 0.65* | 16 ± 1 | 17 ± 1 | 0.17* |
| **Clinical Diagnosis** | |  | 0.95♰ |  |  | 1♰ |
| **Cognitively normal** | 4 (23.5%) | 1 (25%) |  | 0 (0%) | 0 (0%) |  |
| **Mild cognitive impairment** | 13 (76.5%) | 3 (75%) |  | 12 (75%) | 6 (75%) |  |
| **Dementia** | 0 (0%) | 0 (0%) |  | 4 (25%) | 2 (25%) |  |
| **ApoE ε4 count** | |  | 0.33♰ |  |  | 0.14♰ |
| **0** | 11 (64.7%) | 2 (50%) |  | 7 (43.75%) | 3 (37.5%) |  |
| **1** | 3 (17.6%) | 2 (50%) |  | 8 (50%) | 2 (25%) |  |
| **2** | 3 (17.6%) | 0 (0%) |  | 1 (6.25%) | 3 (37.5%) |  |
| **Body mass index (Mean ± SD)** | 26 ± 3 | 26 ± 5 | 0.97* | 27 ± 4 | 26 ± 3 | 0.98* |
| **Systolic blood pressure, mm Hg (Mean ± SD)** | 140 ± 18 | 125 ± 9 | 0.14* | 129 ± 15 | 122 ± 12 | 0.08* |
| **History of hypertension** | 7 (41.2%) | 3 (75%) | 0.22♰ | 9 (56.3%) | 6 (75%) | 0.37♰ |
| **History of diabetes mellitus** | 0 (0%) | 0 (0%) | NA | 1 (6.3%) | 0 (0%) | 0.47♰ |
| **History of heart disease** | 0 (0%) | 0 (0%) | NA | 0 (0%) | 0 (0%) | NA |
| **History of stroke** | 0 (0%) | 0 (0%) | NA | 0 (0%) | 0 (0%) | NA |
| **Use of antihypertensive medication** | 3 (17.6%) | 1 (25%) | 0.74♰ | 2 (12.5%) | 2 (25%) | 0.44♰ |
| **Cigarette smoking status** | 0 (0%) | 0 (0%) | NA | 0 (0%) | 0 (0%) | NA |
| **CSF Phospho-tau/Beta-Amyloid 1-42 Ratio positive status** | 6 (42.9%‡) | 1 (33.3%‡) | 0.76♰ | 8 (61.5%‡‡) | 5 (100%‡‡) | 0.1♰ |
| **CSF Beta-Amyloid 1-42** | 210.9 ± 63.9 | 203 ± 40.5 | 0.77* | 174.8 ± 58.8 | 99.8 ± 19.4 | 0.007* |
| **CSF Phospho-tau** | 27.2 ± 15.1 | 33.3 ±30.2 | 0.86* | 28.5 ± 14.6 | 42.4 ± 17.9 | 0.173* |
| **CSF Total-tau** | 73.9 ± 29 | 58.3 ± 25.1 | 0.59* | 93.8 ± 51.8 | 125.4 ± 87.8 | 0.44* |
| **Time: Plasma to first MRI (years)** | 5.3 ± 0.8 | 5.3 ± 1.1 | 0.76* | Not applicable | Not applicable | - |
| **Time: Plasma to autopsy** | Not applicable | Not applicable | - | 4.04 ± 1.9 | 5.5 ± 0.9 | 0.09* |
| **Change in lobar microbleeds, median (Range)** | 0 (0-1)‡‡‡ | 3 (0-3)‡‡‡ | 0.12* | Not applicable | Not applicable | - |
| **Time: Baseline MRI to follow-up MRI, months** | 42 ± 28‡‡‡ | 60 ± 52‡‡‡ | 0.68* | Not applicable | Not applicable | - |
| **Annual change rate of lobar microbleeds** | 0.01 ± 0.02‡‡‡ | 0.036 ± 0.043‡‡‡ | 0.24* | Not applicable | Not applicable | - |
| **Alzheimer’s Disease – NIA-AA Neuropathological Change level** | | | - |  |  | 0.42♰ |
| **0 (Not AD)** | Not applicable | Not applicable |  | 1 (4.2%) | 0 (0%) |  |
| **1 (Low)** | Not applicable | Not applicable |  | 6 (25%) | 1 (12.5%) |  |
| **2 (Intermediate)** | Not applicable | Not applicable |  | 0 (0%) | 0 (0%) |  |
| **3 (High)** | Not applicable | Not applicable |  | 17 (70.8%) | 7 (87.5%) |  |
| * Mann-Whitney U test. | | | | | | |
| ♰ Chi-square Test. | | | | | | |
| ‡ Percentage among the available cases (available in n=14 and n=3 for CAA- and CAA+, respectively) | | | | | | |
| ‡‡ Percentage among the available cases (available in n=13 and n=5 for CAA- and CAA+, respectively) | | | | | | |
| ‡‡‡ Percentage among the available cases (available in n=14 and n=3 for CAA- and CAA+, respectively) | | | | | | |

Supplementary Table 4. Correlations of lobar microbleed count and neuropathological cerebral amyloid angiopathy severity score with plasma analytes. Only results with p<0.05 in any time points are presented.

|  |  |  | **T0** | | | | **T1** | | | |
| --- | --- | --- | --- | --- | --- | --- | --- | --- | --- | --- |
|  |  |  |  |  | **95% Confidence interval** | |  |  | **95% Confidence interval** | |
|  |  | **Analyte** | **rho** | **p** | **Lower Bound** | **Upper Bound** | **rho** | **p** | **Lower Bound** | **Upper Bound** |
| **MRI - Lobar microbleed count** |  | Angiopoietin-2 | 0.204 | 0.374 | -0.262 | 0.593 | **0.57** | **0.011** | **0.142** | **0.818** |
|  |  | Complement C3 | -0.065 | 0.779 | -0.494 | 0.389 | **-0.503** | **0.028** | **-0.785** | **-0.049** |
|  |  | Epidermal Growth Factor Receptor | -0.358 | 0.111 | -0.691 | 0.101 | **-0.526** | **0.021** | **-0.797** | **-0.08** |
|  |  | FASLG Receptor | **0.488** | **0.025** | **0.058** | **0.765** | 0.387 | 0.101 | -0.096 | 0.723 |
|  |  | Follicle-Stimulating Hormone | -0.185 | 0.421 | -0.58 | 0.28 | **-0.51** | **0.026** | **-0.788** | **-0.058** |
|  |  | Kidney Injury Molecule-1 | **0.519** | **0.016** | **0.099** | **0.782** | **0.472** | **0.041** | **0.008** | **0.769** |
|  |  | Luteinizing Hormone | -0.094 | 0.685 | -0.515 | 0.364 | **-0.534** | **0.019** | **-0.801** | **-0.091** |
|  |  | Neutrophil Gelatinase-Associated Lipocalin | **0.536** | **0.012** | **0.122** | **0.791** | 0.305 | 0.204 | -0.187 | 0.675 |
|  |  | Osteopontin | 0.315 | 0.164 | -0.148 | 0.665 | **0.505** | **0.027** | **0.051** | **0.786** |
|  |  | Prolactin | -0.117 | 0.614 | -0.532 | 0.344 | **-0.466** | **0.045** | **-0.765** | **0** |
|  |  | Receptor for advanced glycosylation end | 0.425 | 0.055 | -0.022 | 0.73 | **0.499** | **0.03** | **0.044** | **0.783** |
|  |  | Tamm-Horsfall Urinary Glycoprotein | **-0.568** | **0.007** | **-0.808** | **-0.168** | -0.381 | 0.107 | -0.719 | 0.103 |
|  |  | Vascular Cell Adhesion Molecule-1 | **0.469** | **0.032** | **0.033** | **0.755** | 0.424 | 0.07 | -0.052 | 0.743 |
|  |  | Vitronectin | **-0.438** | **0.047** | **-0.738** | **0.006** | **-0.484** | **0.036** | **-0.775** | **-0.024** |
| **Neuropathological CAA severity** |  | Alpha-Fetoprotein | -0.379 | 0.068 | -0.685 | 0.042 | **-0.495** | **0.019** | **-0.764** | **-0.079** |
|  |  | Apolipoprotein A-II | **0.522** | **0.009** | **0.138** | **0.77** | 0.414 | 0.055 | -0.023 | 0.718 |
|  |  | Apolipoprotein C-I | **0.502** | **0.012** | **0.112** | **0.758** | 0.136 | 0.546 | -0.315 | 0.537 |
|  |  | Apolipoprotein C-III | **0.624** | **0.001** | **0.283** | **0.825** | **0.463** | **0.03** | **0.038** | **0.746** |
|  |  | Apolipoprotein E | **0.409** | **0.047** | **-0.006** | **0.704** | 0.331 | 0.132 | -0.118 | 0.668 |
|  |  | AXL Receptor Tyrosine Kinase | -0.362 | 0.083 | -0.675 | 0.062 | **-0.497** | **0.019** | **-0.765** | **-0.082** |
|  |  | Clusterin | **0.577** | **0.003** | **0.214** | **0.8** | **0.436** | **0.043** | **0.004** | **0.731** |
|  |  | Complement Factor H | **0.486** | **0.016** | **0.091** | **0.749** | **0.555** | **0.007** | **0.161** | **0.796** |
|  |  | C-peptide | 0.201 | 0.347 | -0.233 | 0.567 | **0.54** | **0.009** | **0.141** | **0.789** |
|  |  | Leptin | 0.2 | 0.348 | -0.233 | 0.567 | **0.448** | **0.036** | **0.02** | **0.738** |
|  |  | Macrophage Migration Inhibitory Factor | -0.057 | 0.792 | -0.46 | 0.366 | **-0.457** | **0.033** | **-0.743** | **-0.03** |
|  |  | Proinsulin-Intact | 0.13 | 0.546 | -0.3 | 0.516 | **0.447** | **0.037** | **0.019** | **0.737** |
|  |  | Testosterone-Total | **-0.497** | **0.014** | **-0.755** | **-0.104** | **-0.559** | **0.007** | **-0.798** | **-0.167** |
|  |  | Thrombomodulin | -0.389 | 0.06 | -0.692 | 0.03 | **-0.467** | **0.029** | **-0.748** | **-0.043** |
|  |  | Transthyretin | **0.428** | **0.037** | **0.017** | **0.715** | 0.204 | 0.363 | -0.251 | 0.585 |

Supplementary Table 5. Area under the curve values of plasma analyte ratios for CAA categories based on either MRI or neuropathological assessments at both time points. Results with false discovery rate (FDR) corrected p <0.05 are marked in bold.

*Abbreviations: AUC, area under the curve; CAA, cerebral amyloid angiopathy; Ang2, Angiopoietin-2; Vitr, Vitronectin; FASLG, FAS Ligand Receptor; KIM1, Kidney Injury Molecule-1; NGAL, Neutrophil Gelatinase-Associated Lipocalin; RAGE, Receptor for advanced glycosylation end; VCAM1, Vascular Cell Adhesion Molecule-1; EGF, Epidermal Growth Factor Receptor; Apo, Apolipoprotein; CLU, Clusterin; ComFH, Complement Factor H; TTR, Transthyretin; Std., standard.*

|  |  | **T0** | | | | | | **T1** | | | | | |
| --- | --- | --- | --- | --- | --- | --- | --- | --- | --- | --- | --- | --- | --- |
|  | Analytes | Area | Std. Error | p | p-FDR | 95% Confidence interval | | Area | Std. Error | p | p-FDR | 95% Confidence interval | |
|  |  |  |  |  |  | Lower Bound | Upper Bound |  |  |  |  | Lower Bound | Upper Bound |
| **CAA – MRI: ≥2 lobar microbleeds** | Ang2/Vitronectin | 0.706 | 0.162 | 0.2 | 0.22 | 0.389 | 1.023 | **0.854** | **0.089** | **0.0001** | **0.0002** | **0.679** | **1.029** |
|  | FASLG/Vitronectin | **0.875** | **0.098** | **0.0001** | **0.001** | **0.684** | **1.066** | **0.979** | **0.031** | **0** | **0** | **0.918** | **1.04** |
|  | KIM1/Vitronectin | 0.721 | 0.122 | 0.07 | 0.09 | 0.482 | 0.96 | 0.688 | 0.156 | 0.23 | 0.25 | 0.382 | 0.993 |
|  | NGAL/Vitronectin | **0.838** | **0.091** | **0.0002** | **0.001** | **0.661** | **1.016** | **0.938** | **0.061** | **0.0000000000005** | **0.000000000002** | **0.819** | **1.056** |
|  | RAGE/Vitronectin | **0.868** | **0.121** | **0.002** | **0.01** | **0.63** | **1.106** | 0.75 | 0.188 | 0.18 | 0.22 | 0.381 | 1.119 |
|  | Osteopontin/Vitronectin | **0.735** | **0.104** | **0.02** | **0.04** | **0.531** | **0.939** | 0.729 | 0.131 | 0.08 | 0.1 | 0.472 | 0.987 |
|  | VCAM1/Vitronectin | **0.809** | **0.096** | **0.001** | **0.004** | **0.62** | **0.997** | **0.813** | **0.098** | **0.001** | **0.002** | **0.621** | **1.004** |
|  | Ang2/EGF | **0.809** | **0.132** | **0.02** | **0.04** | **0.551** | **1.067** | **0.917** | **0.067** | **0.0000000005** | **0.000000002** | **0.786** | **1.048** |
|  | FASLG/EGF | **0.934** | **0.062** | **0.000000000003** | **0.00000000004** | **0.812** | **1.056** | **0.938** | **0.058** | **0.00000000000003** | **0.0000000000002** | **0.825** | **1.05** |
|  | KIM1/EGF | 0.5 | 0.172 | 1 | 1 | 0.162 | 0.838 | 0.5 | 0.219 | 1.00 | 1 | 0.071 | 0.929 |
|  | NGAL/EGF | **0.779** | **0.124** | **0.02** | **0.04** | **0.536** | **1.023** | **0.854** | **0.089** | **0.0001** | **0.00017** | **0.679** | **1.029** |
|  | RAGE/EGF | **0.926** | **0.073** | **0.000000006** | **0.00000004** | **0.783** | **1.07** | 0.792 | 0.14 | 0.04 | 0.052 | 0.518 | 1.066 |
|  | Osteopontin/EGF | 0.721 | 0.122 | 0.07 | 0.09 | 0.481 | 0.96 | **0.833** | **0.094** | **0.0004** | **0.001** | **0.649** | **1.018** |
|  | VCAM1/EGF | 0.706 | 0.144 | 0.15 | 0.18 | 0.423 | 0.989 | **0.792** | **0.112** | **0.01** | **0.01** | **0.572** | **1.011** |
| **CAA – NP: Moderate or severe** | ApoAII/AXL | 0.719 | 0.123 | 0.08 | 0.14 | 0.477 | 0.961 | **0.768** | **0.123** | **0.03** | **0.04** | **0.527** | **1.008** |
|  | ApoCIII/AXL | **0.914** | **0.058** | **0.000000000001** | **0.00000000001** | **0.801** | **1.027** | **0.786** | **0.117** | **0.01** | **0.03** | **0.557** | **1.014** |
|  | CLU/AXL | 0.695 | 0.131 | 0.14 | 0.18 | 0.439 | 0.952 | **0.804** | **0.12** | **0.01** | **0.03** | **0.568** | **1.039** |
|  | ComFH/AXL | **0.789** | **0.107** | **0.01** | **0.02** | **0.579** | **0.999** | **0.848** | **0.086** | **0.00005** | **0.0004** | **0.68** | **1.016** |
|  | ApoCI/AXL | 0.75 | 0.122 | 0.04 | 0.09 | 0.512 | 0.988 | 0.741 | 0.132 | 0.07 | 0.08 | 0.483 | 0.999 |
|  | ApoE/AXL | **0.813** | **0.106** | **0.003** | **0.01** | **0.604** | **1.021** | **0.777** | **0.118** | **0.02** | **0.03** | **0.546** | **1.007** |
|  | C-peptide/AXL | 0.633 | 0.115 | 0.25 | 0.28 | 0.407 | 0.858 | **0.786** | **0.116** | **0.01** | **0.03** | **0.558** | **1.013** |
|  | Proinsulin/AXL | 0.609 | 0.13 | 0.4 | 0.4 | 0.355 | 0.864 | 0.723 | 0.122 | 0.07 | 0.08 | 0.485 | 0.962 |
|  | TTR/AXL | 0.688 | 0.126 | 0.14 | 0.18 | 0.44 | 0.935 | 0.705 | 0.131 | 0.12 | 0.12 | 0.449 | 0.962 |
|  | Leptin/AXL | 0.719 | 0.119 | 0.066 | 0.13 | 0.486 | 0.952 | **0.759** | **0.116** | **0.025** | **0.04** | **0.532** | **0.986** |

Supplementary Table 6. Correlations of annual change rate of lobar microbleed count with plasma analytes.

*Abbreviations: Ang2, Angiopoietin-2; Vitr, Vitronectin; FASLG, FAS Ligand Receptor; KIM1, Kidney Injury Molecule-1; NGAL, Neutrophil Gelatinase-Associated Lipocalin; RAGE, Receptor for advanced glycosylation end; VCAM1, Vascular Cell Adhesion Molecule-1; EGF, Epidermal Growth Factor Receptor; Apo, Apolipoprotein; CLU, Clusterin; ComFH, Complement Factor H; TTR, Transthyretin.*

|  |  | **T0** | | | | | **T1** | | | | |
| --- | --- | --- | --- | --- | --- | --- | --- | --- | --- | --- | --- |
|  |  |  |  |  | **95% Confidence interval** | |  |  |  | **95% Confidence interval** | |
|  | **Analyte** | **Rho** | **p** | **p-FDR** | **Lower Bound** | **Upper Bound** | **Rho** | **p** | **p-FDR** | **Lower Bound** | **Upper Bound** |
| **Ratios - Vitronectin** | Ang2/Vitr | 0.376 | 0.137 | 0.243 | -0.143 | 0.733 | 0.397 | 0.114 | 0.424 | -0.118 | 0.744 |
|  | FASLG/Vitr | 0.562 | 0.019 | 0.081 | 0.096 | 0.826 | 0.19 | 0.465 | 0.543 | -0.333 | 0.624 |
|  | KIM1/Vitr | 0.202 | 0.436 | 0.509 | -0.322 | 0.632 | 0.228 | 0.378 | 0.529 | -0.297 | 0.648 |
|  | NGAL/Vitr | 0.236 | 0.362 | 0.507 | -0.29 | 0.653 | 0.102 | 0.697 | 0.697 | -0.411 | 0.566 |
|  | RAGE/Vitr | 0.005 | 0.986 | 0.986 | -0.489 | 0.496 | 0.301 | 0.24 | 0.466 | -0.224 | 0.691 |
|  | Osteopontin/Vitr | 0.548 | 0.023 | 0.081 | 0.076 | 0.819 | 0.286 | 0.266 | 0.466 | -0.24 | 0.682 |
|  | VCAM1/Vitr | 0.374 | 0.139 | 0.243 | -0.145 | 0.732 | 0.391 | 0.121 | 0.424 | -0.125 | 0.741 |
| **Ratios - EGF** | Ang2/EGF | 0.513 | 0.035 | 0.245 | 0.027 | 0.803 | 0.507 | 0.038 | 0.203 | 0.019 | 0.8 |
|  | FASLG/EGF | 0.45 | 0.07 | 0.245 | -0.054 | 0.772 | 0.467 | 0.059 | 0.203 | -0.033 | 0.78 |
|  | KIM1/EGF | -0.067 | 0.798 | 0.798 | -0.542 | 0.44 | -0.087 | 0.741 | 0.741 | -0.555 | 0.424 |
|  | NGAL/EGF | 0.269 | 0.296 | 0.484 | -0.257 | 0.673 | 0.423 | 0.091 | 0.203 | -0.088 | 0.758 |
|  | RAGE/EGF | 0.1 | 0.701 | 0.798 | -0.412 | 0.565 | 0.346 | 0.174 | 0.203 | -0.177 | 0.716 |
|  | Osteopontin/EGF | 0.405 | 0.107 | 0.250 | -0.109 | 0.748 | 0.394 | 0.118 | 0.203 | -0.123 | 0.742 |
|  | VCAM1/EGF | 0.244 | 0.346 | 0.484 | -0.283 | 0.657 | 0.361 | 0.155 | 0.203 | -0.16 | 0.724 |
| **Ratios - AXL** | ApoAII/AXL | -0.288 | 0.263 | 0.557 | -0.683 | 0.239 | -0.295 | 0.25 | 0.380 | -0.688 | 0.231 |
|  | ApoCIII/AXL | -0.274 | 0.287 | 0.557 | -0.675 | 0.253 | -0.175 | 0.502 | 0.558 | -0.615 | 0.347 |
|  | CLU/AXL | -0.25 | 0.334 | 0.557 | -0.661 | 0.277 | -0.253 | 0.328 | 0.410 | -0.663 | 0.274 |
|  | ComFH/AXL | -0.102 | 0.697 | 0.774 | -0.566 | 0.411 | -0.018 | 0.945 | 0.945 | -0.506 | 0.479 |
|  | ApoCI/AXL | -0.359 | 0.157 | 0.557 | -0.724 | 0.162 | -0.336 | 0.187 | 0.380 | -0.711 | 0.187 |
|  | ApoE/AXL | -0.394 | 0.117 | 0.557 | -0.743 | 0.122 | -0.531 | 0.028 | 0.280 | -0.811 | -0.052 |
|  | C-peptide/AXL | -0.041 | 0.876 | 0.876 | -0.523 | 0.461 | -0.419 | 0.094 | 0.380 | -0.755 | 0.093 |
|  | Proinsulin/AXL | -0.102 | 0.697 | 0.774 | -0.566 | 0.411 | -0.359 | 0.157 | 0.380 | -0.724 | 0.162 |
|  | TTR/AXL | -0.14 | 0.592 | 0.774 | -0.592 | 0.379 | -0.288 | 0.263 | 0.380 | -0.683 | 0.239 |
|  | Leptin/AXL | -0.329 | 0.198 | 0.557 | -0.707 | 0.195 | -0.286 | 0.266 | 0.380 | -0.682 | 0.24 |

Supplementary Figure 1. Flowchart of the initial number of participants and those excluded.

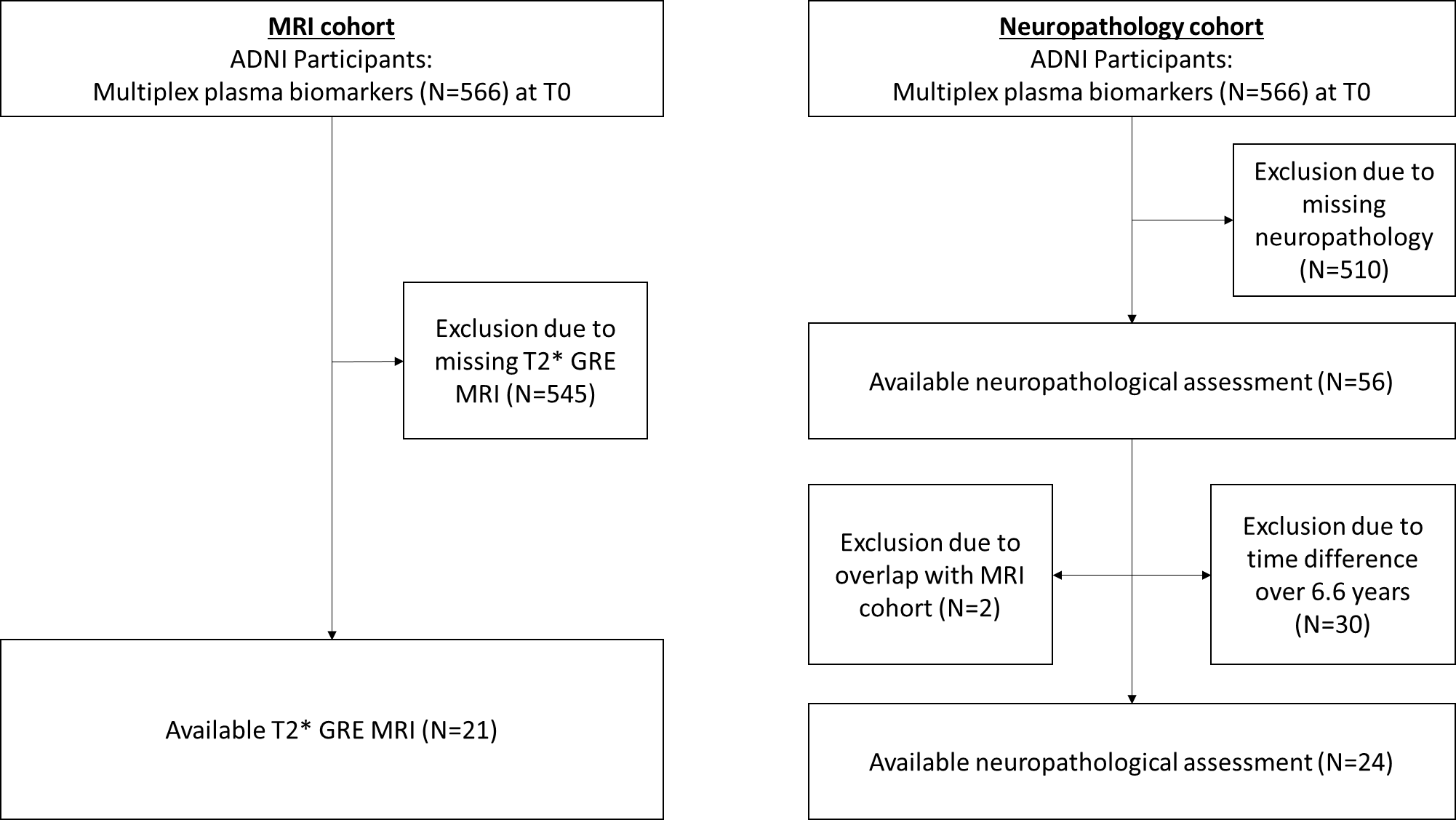

Figure legends (separately provided figures):

Supplementary Figure 2. Scatter plots between ratios of plasma analytes and CAA proxy measures among the study groups and timepoints. *Abbreviations: NPAMY. neuropathological severity scale for CAA.*

Supplementary Figure 3. Scatter plots between annual change rate of lobar microbleed count and plasma analytes (A. FASLG/Vitronectin, B. Osteopontin/Vitronectin, C. Ang2/EGF and D. ApoE/AXL) for both time points*.*
