## Supplementary figures and images for "An explorative analysis of plasma biomarkers associated with cerebral amyloid angiopathy"

### Supplementary Figure 2

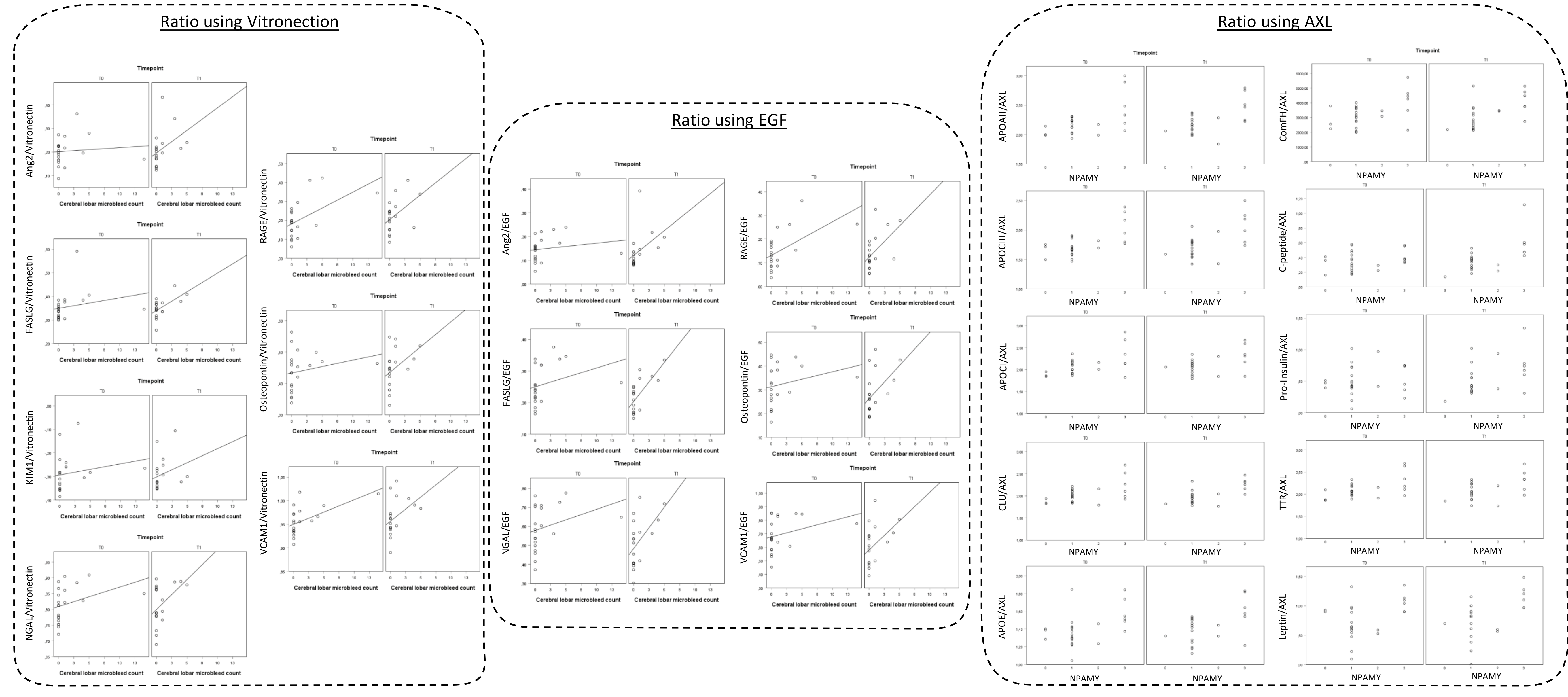

### Supplementary Figure 3

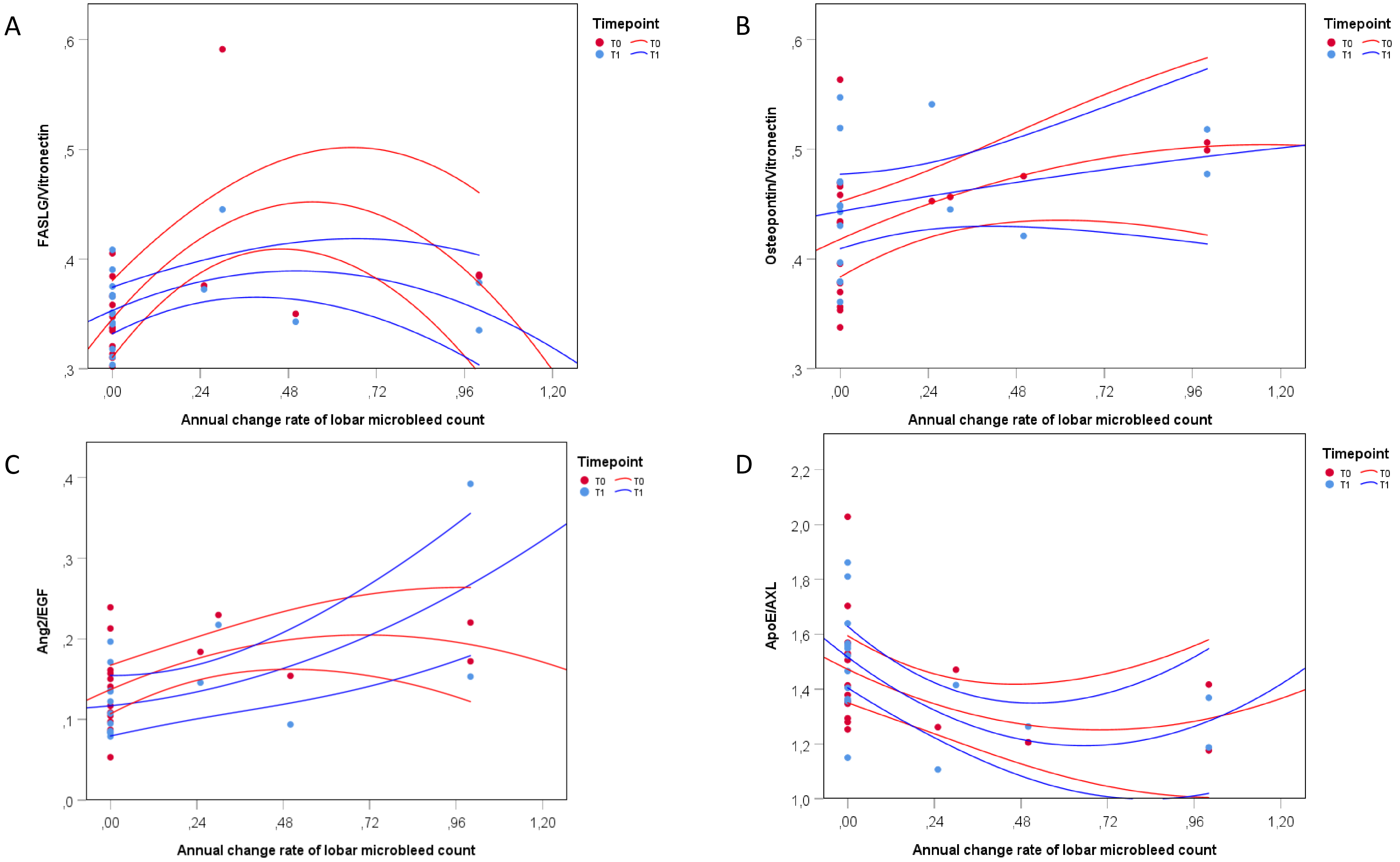
